# Recent Advances in the Diagnosis of Autoimmune Inner Ear Disease: A Scoping Review

**DOI:** 10.1101/2025.01.22.25320943

**Authors:** Jialin Liu, Changyu Wang, Siru Liu

## Abstract

Autoimmune inner ear disease (AIED) is a rare cause of sensorineural hearing loss, typically characterized by progressive bilateral hearing loss and vestibular symptoms. Diagnosing AIED is challenging due to the lack of specific biomarkers and its overlapping clinical features with other causes of hearing loss. This scoping review examines the recent advancements in the diagnostic approaches for AIED from 2020 to 2024, focusing on studies indexed in SCI, MEDLINE, and INSPEC. The review highlights progress in laboratory tests, imaging techniques, and genetic assessments, and discusses the implications of these advances in improving early diagnosis and treatment outcomes.

## Introduction

Autoimmune inner ear disease (AIED) is a rare and complex disorder characterized by progressive and often fluctuating sensorineural hearing loss (SNHL) [1]. The incidence of AIED is estimated to be less than 5 per 100,000 people per year, with an estimated incidence of 45,000 in the United States [4]. There are two forms of AIED, primary and secondary [5]. Primary AIED involves pathology confined to the inner ear. Secondary AIED occurs in about one-third of cases. It is associated with systemic autoimmune conditions. Examples of these conditions include systemic lupus erythematosus (SLE), rheumatoid arthritis (RA), and autoimmune thyroid disease [2,3].

The hallmark symptoms of AIED include bilateral SNHL, tinnitus, and occasionally vestibular disturbances such as vertigo [6]. The condition arises from an aberrant immune response, where the body’s immune system mistakenly targets the inner ear structures [7]. While AIED can occur independently, its association with systemic autoimmune diseases and the lack of specific diagnostic tests complicate its identification and management [6,8].

Historically, the diagnosis of AIED relied on clinical criteria, primarily through the exclusion of other potential causes of hearing loss [6,8,9]. However, the absence of reliable, disease-specific diagnostic markers has often led to delays in diagnosis and treatment, contributing to irreversible hearing damage and poorer patient outcomes. Recent advancements in genetic testing, biomarkers, imaging, and serologic assays show promise in diagnostics but require further validation for AIED [10-15].

This scoping review aims to synthesize the latest advancements in AIED diagnostics, focusing on key studies published between 2020 and 2024, with the goal of providing an updated understanding of diagnostic strategies and their implications for clinical practice.

## Methods

### Search Strategy

A systematic search of the literature was conducted in the following databases: Web of Science Core Collection, MEDLINE, and INSPEC, covering articles published between January 2020 and December 2024. The search terms included “autoimmune inner ear disease” AND “diagnosis,” combined with Boolean operators to expand or refine the search scope. Synonyms and related terms were also included to ensure comprehensive coverage. For example, “AIED” was combined with “diagnostic advancements,” “imaging techniques,” and “genetic markers.” To identify relevant studies, two authors independently screened titles and abstracts based on predefined inclusion criteria. Discrepancies in selection were resolved through discussion or consultation with a third author. The inclusion criteria were as follows: 1) Peer-reviewed articles published between January 2020 and December 2024; 2) Studies focusing on diagnostic methodologies, including imaging techniques, genetic markers, or advanced tests for AIED; 3) Articles published in English. Studies were excluded: 1) Did not focus on AIED or its diagnosis; 2) Were review articles, letters, abstracts, or conference proceedings; 3) Were duplicates or otherwise irrelevant to the research question.

### Data Extraction and Synthesis

Data extraction was conducted systematically to capture key details of the diagnostic methods explored, including their respective sensitivities, specificities, and clinical impacts. Additionally, general characteristics of the studies, such as sample size, population demographics, study design, and methodologies employed, were meticulously recorded. The extraction process utilized a standardized data extraction template to ensure uniformity and minimize potential bias.

A narrative synthesis approach was adopted to integrate and summarize the findings. This method facilitated the identification of common trends, methodological strengths and weaknesses, and research gaps within the literature. Studies were categorized based on diagnostic modalities, such as imaging techniques, genetic markers, and advanced diagnostic tests, to provide a structured analysis. Where possible, thematic analysis was performed to highlight recurring patterns and unique insights.

## Results

### Literature Search and Selection Process

Based on the search strategy, 67 documents were initially retrieved. After a thorough screening process, including the assessment of literature type, titles, abstracts, removal of duplicates, and full-text review, 5 studies were selected for inclusion in the review (Figure 1).

**Figure 1.**
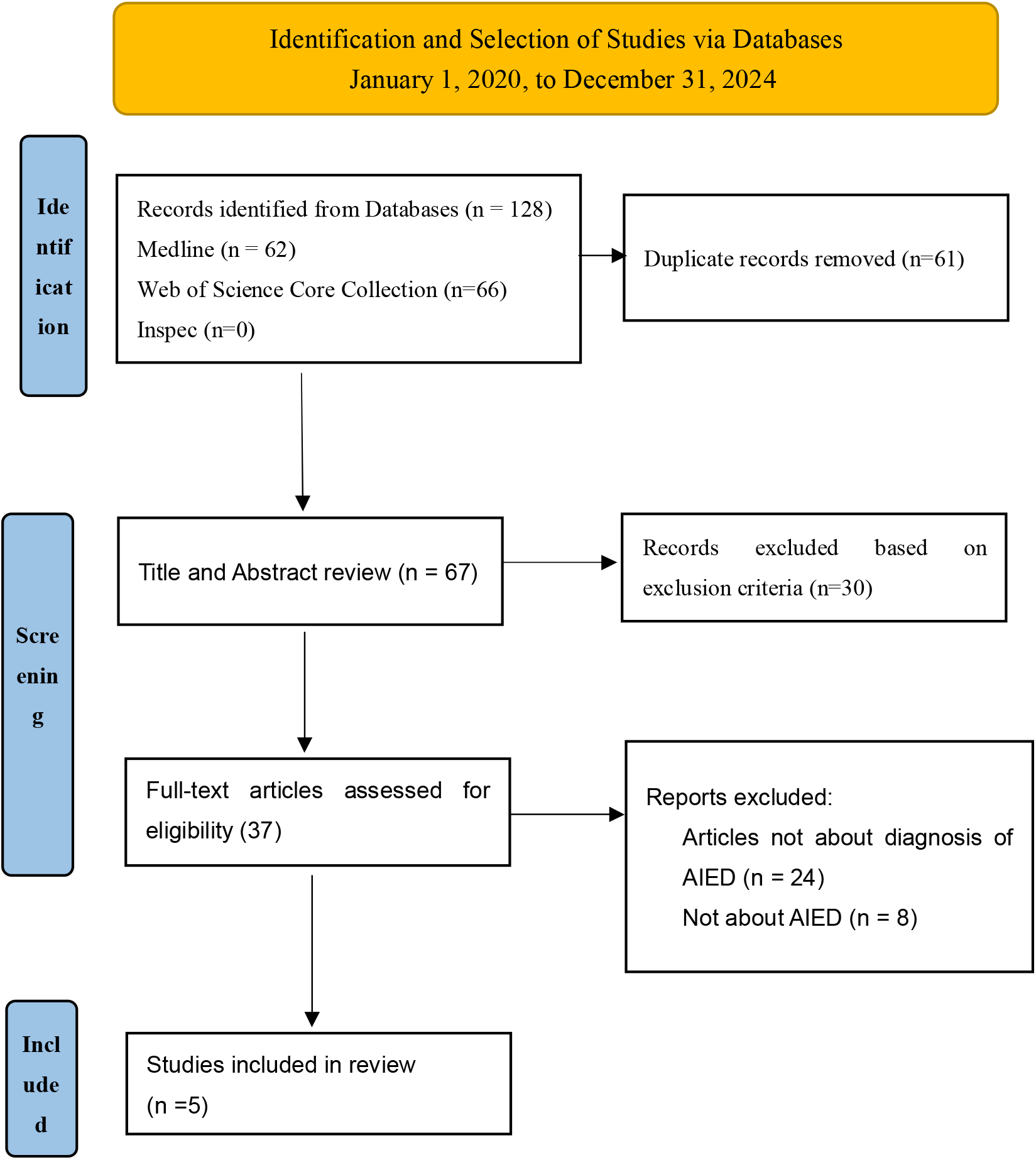
PRISMA flow diagram of the studies selection.

### Characteristics of Included Studies

A total of 5 studies were included in this review, comprising 1 case-control study and 4 retrospective studies, with a combined sample size of 163 patients. Of these, 110 were females (67.5%). The age range of the patients was 24 to 81 years. The included studies investigated various aspects of AIED, including: diagnostic criteria, audiometric patterns, the role of antiphospholipid antibodies (aPL) in AIED, and clinical features of AIED. Additionally, one study explored the use of MRI as a diagnostic tool to differentiate AIED from other causes of SNHL (Table 1).

**Table 1.**
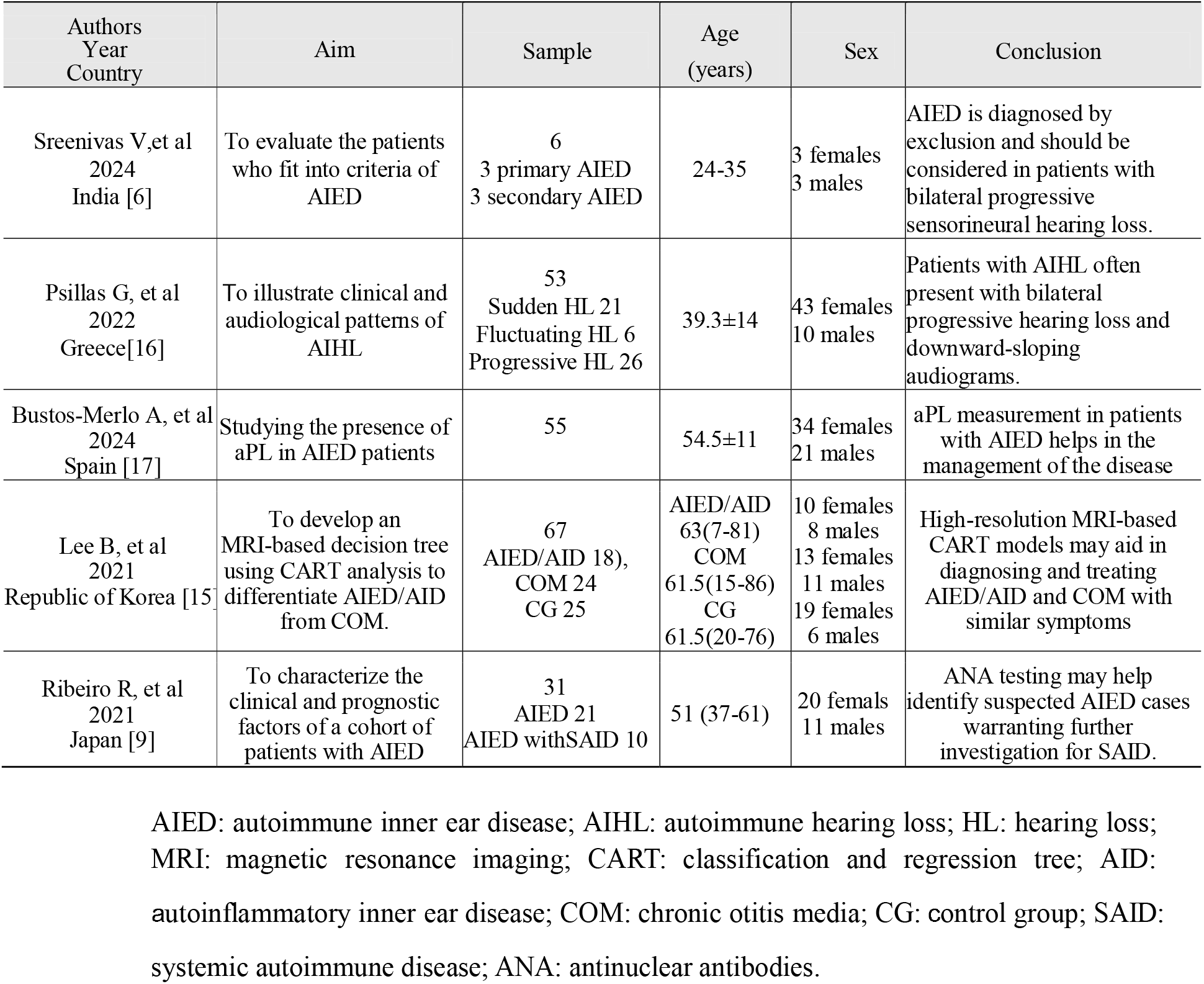
Characteristics of studies included from full text screening.

## Discussion

As there is no gold standard for the diagnosis of AIED, diagnosis requires a comprehensive consideration of the patient’s condition. AIED is slightly more common among females than males, consistent with the higher prevalence of autoimmune diseases in females. The typical age of onset ranges is between 20 and 50 years [18]. It mainly involves clinical signs and symptoms, laboratory tests, imaging and steroid treatment. The following are key points to note in the diagnosis of AIED.

### Clinical evaluation

The diagnosis of AIED relies on excluding other causes of SNHL and recognizing its distinctive clinical features. The evaluation encompasses both auditory and systemic manifestations, focusing on the following aspects [6,7,8,19]:

1. Onset and Progression of Hearing Loss: AIED typically presents as rapidly progressive or fluctuating bilateral SNHL, though it may occasionally begin unilaterally. Hearing loss is usually progressive, with a reduction of at least 30 dB at one or more frequencies.
2. Associated Aural Symptoms: Common symptoms include a sensation of ear fullness, tinnitus and episodic or persistent vertigo, reflecting auditory and vestibular system involvement.
3. Predisposing Factors and History: Identifying risk factors such as noise exposure, ototoxic medications (e.g., Aminoglycosides or chemotherapy), head trauma, prior ear surgeries, meningitis, or a family history of hearing loss is crucial.
4. Systemic Autoimmune Features: AIED may occur in isolation or as part of a systemic autoimmune disease. Evaluation for systemic symptoms such as joint pain, skin rashes, swelling, dry eyes or mouth, glandular swelling and dysphagia is essential for early detection.
5. Ear and Vestibular Examination: A thorough examination of the external and middle ear is essential to rule out other potential causes of hearing loss. Vestibular function assessments can identify inner ear involvement.
6. Audiometric Testing: A comprehensive audiometric evaluation, encompassing pure-tone audiometry and speech audiometry, is crucial for accurately documenting the degree and type of hearing loss, as well as for monitoring disease progression and treatment response.

Given the autoimmune basis of AIED, a detailed patient history and thorough physical examination are indispensable for identifying potential autoimmune triggers or systemic involvement. These steps, combined with the careful exclusion of other etiologies, form the foundation for an accurate diagnosis and a tailored treatment strategy.

### Laboratory Examination

Laboratory tests are essential for supporting the diagnosis of AIED by identifying immune system abnormalities and ruling out alternative causes of SNHL. The recommended laboratory examinations for AIED include [6, 20,21]:

1. Complete Blood Count (CBC) with Differential: Abnormal white blood cell counts or the presence of eosinophilia may indicate systemic inflammation or autoimmune involvement.
2. Erythrocyte Sedimentation Rate (ESR): Elevated ESR suggests an inflammatory or autoimmune process commonly associated with AIED.
3. Antinuclear Antibody (ANA): A positive ANA test may indicate systemic autoimmune diseases, such as lupus, that can present with inner ear symptoms.
4. Immunological Profile: Broader immune activity is evaluated, including levels of immunoglobulins (IgG, IgA, IgM) and complement proteins (C3, C4), which may reveal irregularities in autoimmune conditions [22].
5. Anti-HSP70 Antibody: Positive anti-HSP70 antibodies are suggestive of autoimmune activity targeting the inner ear. One study reported anti-HSP70 with a sensitivity of 79.07%, specificity of 100%, positive predictive value of 100%, negative predictive value of 76.92%, and overall diagnostic accuracy of 87.67% in AIED cases [23]. However, a systematic review concluded that evidence is insufficient to recommend HSP70 autoantibody testing as a routine diagnostic tool for immune-mediated inner ear disease [24].
6. Antiphospholipid Antibodies (aPL): Positive results may indicate antiphospholipid syndrome, which is associated with vascular complications and SNHL [17].

These laboratory tests complement clinical evaluations by confirming autoimmune activity and excluding infectious or other non-immune causes of hearing loss. Although no single test is definitive for the diagnosis of AIED, a constellation of findings, including elevated ESR, positive anti-HSP70 antibodies, and aPL, can strengthen the diagnosis. Laboratory results should be interpreted alongside clinical findings and audiometric evaluations to guide diagnosis and treatment [25]. Current routine immunological laboratory tests, including antinuclear, antineutrophil cytoplasmic, antiendothelial cell, antiphospholipid/anticardiolipin, and antithyroid antibodies, are also recommended when assessing patients with suspected immune-mediated inner ear disorders [26].

### Imaging Examinations

Advancements in imaging technology have significantly improved the evaluation of AIED’s effects on the inner ear. Magnetic resonance imaging (MRI) and computed tomography (CT) are indispensable diagnostic tools, especially when clinical symptoms overlap with other causes of hearing loss. Although cochlear enhancement on MRI is not specific to AIED, it may indicate inner ear inflammation consistent with the disease. Intratympanic gadolinium-enhanced MRI has shown potential in visualizing inner ear structures and detecting inflammatory changes [27,28]. Additionally, gadolinium (Gd)-enhanced MRI using 3D fluid-attenuated inversion recovery (FLAIR) imaging is particularly useful when inner ear pathology is suspected [29]. These imaging techniques not only enhance diagnostic precision but also help differentiate AIED from other conditions, thereby facilitating earlier detection and tailored interventions. Incorporating these modalities into the diagnostic workflow allows clinicians to identify AIED with greater accuracy and implement targeted treatments, ultimately improving patient outcomes.

### Genetic Testing

Genetic testing has emerged as a valuable tool for uncovering susceptibility factors in AIED, particularly in cases with familial clustering or idiopathic origins. Recent studies have identified specific genetic polymorphisms that may increase predisposition to AIED [30]. Notably, certain human leukocyte antigen (HLA) haplotypes, including HLA-B27, B35, B51, C4, C7, and A1-B8-DR3, have been investigated for their potential roles as prognostic markers in AIED-related hearing loss [31-37]. Additionally, genetic polymorphisms in interleukin-1 receptor (IL-1R) genes have been associated with increased susceptibility to sudden SNHL [38]. By identifying genetic predispositions, genetic testing offers valuable insights into disease mechanisms, aids in risk stratification, and informs personalized treatment strategies. These advancements underscore the critical role of genetic research in improving the understanding and management of AIED, paving the way for more targeted and effective therapeutic approaches.

### Diagnostic Therapy

The response to steroid therapy is considered an essential clinical criterion for diagnosing AIED [39]. Corticosteroids, the mainstay of treatment, are often used as both a therapeutic and diagnostic tool. A positive response, typically characterized by improved hearing and reduced vestibular symptoms, can help confirm the diagnosis. However, variability in treatment response and the risk of relapse upon tapering steroids present challenges in relying solely on this approach for diagnosis [6]. Some studies have shown steroid treatment to be 14-70% effective [16, 41-43].

### Future Directions and limitation

Despite recent advances, the diagnosis of AIED remains challenging. Future research should prioritize the identification of more specific biomarkers and the development of standardized diagnostic criteria. The integration of genomic, proteomic, and metabolomic studies may offer deeper insights into the pathophysiology of AIED and facilitate the discovery of novel diagnostic markers. Additionally, advancing imaging techniques and exploring the role of immune profiling could further enhance diagnostic accuracy. Collaborative, multicenter studies are essential to validate these approaches and establish consensus guidelines for the diagnosis and management of AIED.

This review acknowledges several limitations. Firstly, it is subject to language bias, as only peer-reviewed literature published in English was included. This restriction may result in information and selection bias, potentially omitting valuable insights or alternative perspectives from non-English studies. Secondly, the rarity of AIED contributes to the limited number of included studies, which may affect the generalizability and robustness of the findings presented in this review.

## Conclusion

Recent advancements in the diagnosis of AIED have enhanced the ability to identify this rare disease with greater accuracy and at earlier stages. Laboratory diagnostics, advanced imaging techniques, and genetic testing have collectively contributed to a deeper understanding of AIED’s pathophysiology. Despite these developments, critical challenges persist, including the need for more specific biomarkers and the validation of imaging modalities. Future research should prioritize large-scale, multicenter studies to evaluate the clinical utility of emerging diagnostic approaches and establish standardized protocols for diagnosing AIED. Continued progress in these areas will likely facilitate early detection and enable the implementation of personalized treatment strategies, ultimately improving patient outcomes and preventing further auditory and vestibular deterioration.

## Data Availability

All data produced in the present study are available upon reasonable request to the authors

